# A PRACTICAL MODEL FOR EARLY IDENTIFICATION OF PROSPECTIVE HIGH NEED HIGH COST PATIENTS

**DOI:** 10.1101/2022.02.04.22270056

**Authors:** Avivit Golan Cohen, Shlomo Vinker, Adi Isaacson, Eva Avramovich, Eugene Merzon

**Author notes:** **Corresponding author: <u>Dr. Avivit Golan-Cohen</u>**, 23 Shprinzak St., Tel Aviv, Israel. <u>Ethics approval</u>: The study protocol was approved by the Shamir Medical Center Review Board and the Research Committee of LHS (ID number 0235-18-ASAF,Date: 9.10.2018). <u>Consent for publication</u>: Not applicable. <u>Availability of data and materials</u>: The datasets analyzed during the current study are not publicly available because it is a business Information, but are available from the corresponding author on reasonable request. <u>Competing interests</u>: The authors declare that they have no competing interests. <u>Funding</u>: The authors declare that there was no funding in this research. **Authors’ contribution:** AVG: contributing the research questions and study design, literature search, data collection, analysis and interpretation, writing the manuscript. SV: contributing the research questions and study design, drafting the manuscript, analysis and interpretation. AI: literature search, data analysis and interpretation, writing the manuscript. EA: data interpretation, editing the manuscript. EM: contributing the research questions and study design, data collection, analysis and interpretation, drafting the manuscript.

## Abstract

**Background:** High Need-High Cost (HNHC) patients are those who experience poor health outcomes and high health care costs. Early identification may improve outcomes and lower costs.

**Aim:** Development of a model using retrospective data to identify patients at risk for becoming HNHC patients, in order to efficaciously plan interventions.

**Methods:** Data from a large Israeli Health Maintenance Organization (HMO) that includes 488,615 clients above the age of 21 were examined. Multivariate linear regression models were developed using 2012–2016 health expenditure as a dependent variable.

**Results:** The number of yearly purchases of medications for chronic disorders, yearly outpatient visits, yearly emergency department and hospital admissions and the last measured HgA1c level were highly predictive of increased expenditure over a five-year period. Each of these indicators has a different coefficient of influence.

**Conclusions:** We developed a predictive model, based on easily obtained data from electronic medical records that enabled us to identify a population at risk for becoming HNHC in the next five years, a time window allows for intervention. Further research is needed to evaluate whether this is an early enough stage to implement pro-active intervention in the primary care setting.

**Trial registration:** retrospectively registered.

**HIGHLIGHTS:** In this study, we developed a numerical point system calculator, to indicate a risk score for health deterioration within 5 years of patients, by using numerical indicators existing in standard EMR data.

The indicators introduced into this calculated risk can guide healthcare providers to the needed areas of intervention. The display of indicators also promotes optimization of care management and continuity of care.

This risk score is expected to focus the attention of primary care teams on the population that will benefit most from it, as well as to evaluate the effectiveness of specific interventions.

## INTRODUCTION

Coping with the rising prevalence of chronic diseases is one of the major challenges facing health-care systems throughout the world. The increase in both, longevity and disease complexity, has led to a growing number of patients with multiple concurrent morbidities; ranging from 50% to 98% of people older than 65 years in various studies [1-2]. These comorbidities have complex interactions that can influence the progression and management of both disorders [1]. From the economic point of view, complex co-morbidities translate to tripling the national expenditure on health per capita, and it is expected to continue to rise [2]. The increasing burden on resources is compelling healthcare systems to seek ways to ease the burden.

Analyses of health expenditures worldwide reveals a pattern; approximately 5% of the population incurs roughly 50% of all national health care costs [3-5]. Therefore, in an attempt to stem the tide of rising costs, the focus has been on this 5% of the population. It was assumed that a comprehensive approach would reduce multi-morbidity impact, thus resulting in reduced expenditure [6].

Effective management requires a practical algorithm to identify which patients are more likely to deteriorate and to generate substantial future health expenditures. Identification of “complex patients” would be effective and can lead to reduced health care costs only if it occurs in a timely manner. The treatment plan can then be adjusted to impact the course of illness [4]. Originally, researchers focused on patients with specific single diseases known to have major health repercussions (such as CHF or diabetes) or patients with multi-morbidity [1, 4]. Many studies have been conducted on modeling healthcare costs data as an estimator of medical complexity [7-10]. However, when the identified patients presented medically complex conditions, the ability to predict who would deteriorate and become HNHC was limited [11-12].

In 1987, the Charlson Comorbidity Index was developed to predict the risk of death within a year of hospitalization based on the number of chronic diseases [7]. Charlson et al. used 19 categories of diseases, each with a score of 1 to 6. Higher overall calculated score led to a greater chance of dying in the following year. Over the years, the index underwent modifications and in 2008 was validated and adjusted so that it could predict a future increase in medical expenditure [8]. Several additional models have been developed (chronic illness and disability payment system [CDPS], diagnostic cost groups [DCGs], RxGroups, episode risk groups [ERGs]) [10] are among the most quoted. These models use medical insurance claims to generate the main predictor variables [9]. Some of these models predict patient’s chances of dying, some the odds of hospitalization and others predict future total healthcare costs. It turns out that not all patients with multiple chronic diseases deteriorate in a manner that would increase their medical expenditures. Complexity measured only by a quantitative count of chronic diseases does not produce an accurate and useful prediction of the individual patient that may benefit from a proactive intervention [5].

In recent years, the Commonwealth Fund and others have brought to the forefront the role of functional and psychosocial difficulties associated with significant risk factors for future health deterioration [1, 3-5, 11-12]. They found that patients with three or more chronic diseases are more likely to deteriorate if they have functional limitations, poor disease self-management, economic problems, or lack a support system, resulting in fragmentation and poor coordination of care [13-14] and increased pressure on primary care professionals (PCPs) [14]. Following these new insights, the terminology pertaining to the 5% of the population that incurs 50% of health expenditure has been changed to “High Need High Cost” (HNHC) patients. The term reflects the understanding that the management of all complex patient needs, including those beyond the medical disorders, is responsible for the high cost.

An effective definition of the intervention group [18-19] needs to identify not only the patients at risk of deterioration, but within this group, those who would benefit from the intervention. Therefore, the indicators chosen should reflect not only a patient’s current state, but through continuous monitoring, they should also reflect any future changes incurred through interventions. Monitoring enables evaluation of the effectiveness of the intervention, and can potentially motivate both the staff and the patients.

Of the published accepted characteristics of HNHC, we chose indicators associated with high health expenditures such as - clinical parameters (e.g. certain diseases or treatments or lab results that indicate an unbalanced disease state), health resources utilization (e.g. multiple inpatient hospitalizations or multiple physician visits) or patient’s executive functioning (e.g. compliance to medication use or to follow-up visits of physician) [10].

We excluded indicators that did not meet the SMART model (Specific, Measurable, Attainable/Achievable, Realistic/Relevant, and Time-Bound) [20] which is the most common model for the examination of the compatibility of selected indicators. According to the SMART model, indicators should be:

1. Specific – accurately describe the value we want to measure.
2. Measurable – return the same value every time it is measured under the same conditions.
3. Attainable – easy to extract from existing data.
4. Relevant – directly connected to the desired intervention and influenced by it.
5. Time-bound – changes can be tracked over a period of time.

Exclusion of indicators that cannot be influenced or changed is important because their inclusion would skew the results and create a score that is not sensitive to changes. A good example of this is age - an eminent risk factor for deterioration that would mask improvements in any other indicator.

The objective of our study was to formulate a predictive model based on the selected simple numerical indicators from the electronic medical records (EMRs), which would allow us to identify a population at risk for becoming HNHC, at a stage with potential for pro-active intervention in the primary care setting.

## METHODS

Leumit Health Services (LHS) is an HMO in Israel serving about 720,000 members. LHS has a comprehensive computerized database continuously updated with patients’ demographics, medical encounters, laboratory tests and medication prescriptions, and healthcare services utilization.

### Study period and population

#### Study period

our study was conducted retrospectively on records from 2011 until 2016.

**Study population** consisted of all LHS members over the age of 21 at the time of the study period - 488,615 patients.

#### Exclusion criteria

patients who, at the beginning of the study, were diagnosed with cancer, patients requiring dialysis, and patients with diseases defined by the Israeli Ministry of Health as “costly” and covered with a special budget – Thalassemia major, Gaucher, Hemophilia and AIDS.

Patients who died or left the HMO during the study period were also excluded. After these exclusions, the data of 460,912 patients was analyzed.

Total expenditures for 2011 for each patient served as the baseline against which the change in expenditure was measured during the evaluation period. Total medical costs were calculated for each patient for each year from 2012 to 2016, including: medications, hospital admissions, emergency department (ED) visits, office-based physician visits (primary care and consultants),hospital outpatient visits, ambulatory medical services (laboratory and imaging procedures, physical and occupational therapy, ambulances, etc.) and medical devices (such as insulin pumps and supplies, glucometers, orthopedic assistance devices, etc.). These were obtained directly from the billing database of LHS.

Data regarding chronic medication consumption was drawn from the LHS pharmacy claims, based on the Anatomical Therapeutic Chemical (ATC) classification system. A medication was considered chronic if the patient purchased at least 3 prescriptions during the 180 days preceding the index date.

The number of hospital admissions and ER visits was calculated according to hospital claims. Data on visits to primary care physicians and community consultants were obtained from the LHS EMR. Only face to face visits in an LHS ambulatory setting were included.

Uncontrolled diabetes mellitus (DM) was defined according to the last measured HgA1c level result (above 8%) in 2011 and reflected poor glycemic control.

Multivariate linear regression models were created using the following formula on the 2012– 2016 data and the stepwise approach to locate the most influential variables. In order to improve the statistical test, a deviation of at least 5 standard deviations was filtered out.

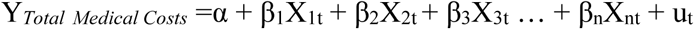

Y- is the dependent (explained) variable.

α – is the constant alpha. In our case it reflects the “minimum” average expenditure given the other variables.

X_n_ – are the independent (explanatory) variables X_1_, X_2_, X_3_ … X_n_

β – is the coefficient for each independent (explanatory) variable β_1_, β_2_, β_3_,.. etc

u – is a random interference, the part that is not explained by the model.

Regression analysis calculated all independent (X) variables in the analysis as numerical. Additionally, we used dummy variables and when the coefficient changed mid-regression (e.g. for HgA1C), we utilized the piecewise approach.

The ***p-value*** of each variable and the adjusted ***R*-*squared*** values were used to test all possible linear regression models to predict total medical costs in combination with the other available independent variables. ***R*-*squared*** *is* the proportion of variability in a data set that provides a measure of how well the future outcome variable (total medical costs) is likely to be predicted by the independent continuous, ordinary and dummy variables in our model. The Kolmogorov– Smirnov test was used for the goodness of fit test. Assumptions were two sided with an α of 0.05. Statistical analysis was performed using STATA 12.0.

The study had no external funding source.

## RESULTS

Baseline study population characteristics are presented in **Table 1**. As expected, age was a remarkable predictor for future cost, both as a categorical (**Table 2**) and as a continuous (**Table 3**) variable. Although age was highly correlated with future health expenditure we decided to exclude it from the model. After exclusion of age R squared remained almost the same. After inclusion age as a categorical and not a continuous variable the result didn’t changed.

**Table 1.**
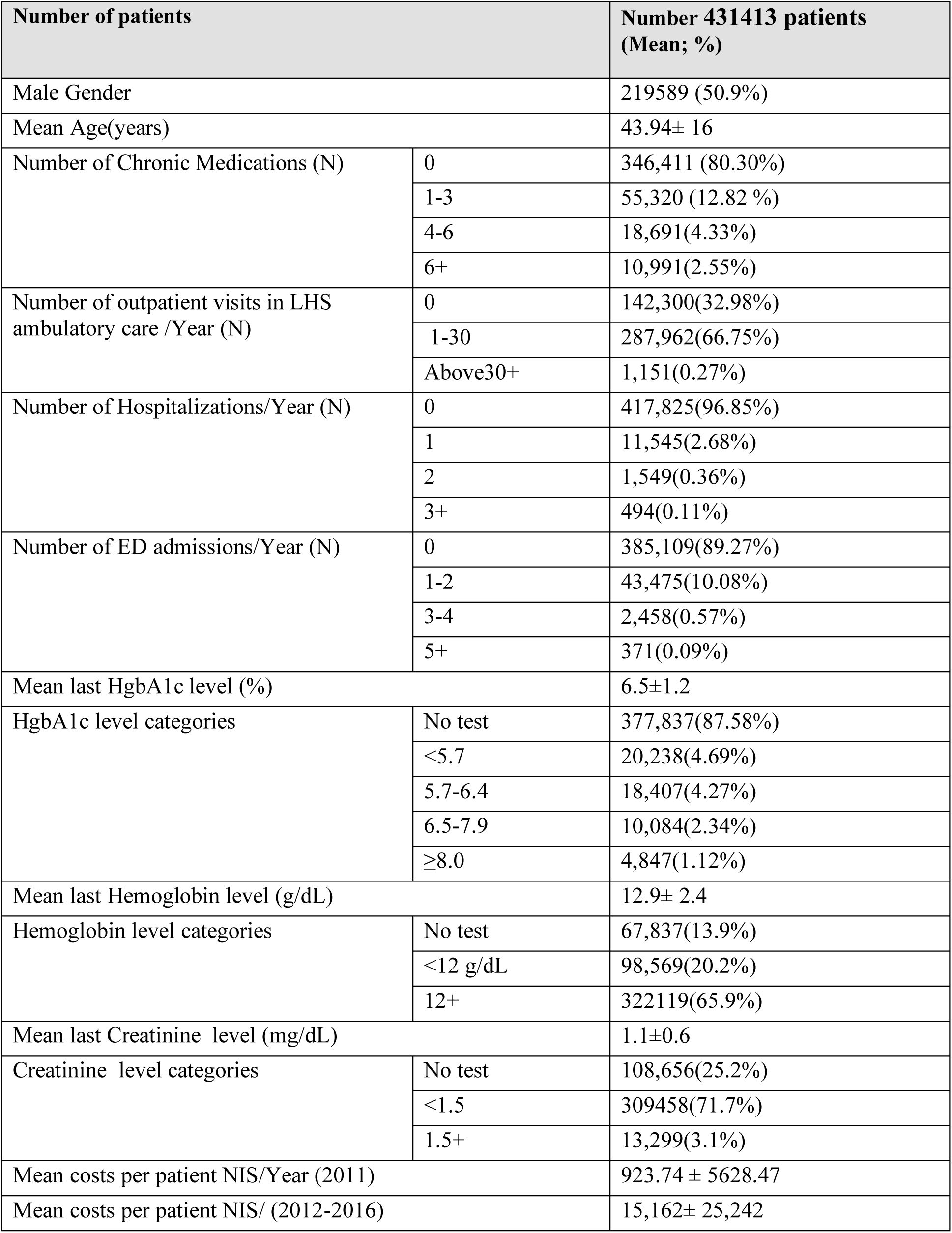
Population characteristics at baseline, 2011.

**Table 2:** Distributions of total future medical costs.

| Variable |  | Mean total costs per patient NIS (2012-2016) |
| --- | --- | --- |
| Age categories | ≤30 | 2132.37 ± 1307.56 |
|  | 31-60 | 4442.39 ± 2089.73 |
|  | 61+ | 15757.44 ± 4113.1 |
| Number of Chronic Medications (N) | 0 | 3280.535±164.22 |
|  | 1-3 | 10591.11±345.77 |
|  | 4-6 | 20818.11± 4641.99 |
|  | 6+ | 36311.93± 6194.6 |
| Number of outpatient visits in LHS ambulatory care /Year (N) | 0 | 1654.91±1308.23 |
|  | 1-30 | 7801.157±2822.03 |
|  | Above30+ | 24846.91±4984.58 |
| Number of Hospitalizations/Year (N) | 0 | 5273.59 ± 2263.19 |
|  | 1 | 19844.25± 4983.66 |
|  | 2 | 33852.15±7267.28 |
|  | 3+ | 51725.59±10138.6 |
| Number of ED admissions/Year (N) | 0 | 5310.188 ± 23536.83 |
|  | 1-2 | 9856.694 ± 31296.62 |
|  | 3-4 | 12238.45± 32466.32 |
|  | 5+ | 18669.32 ± 53509.71 |
| Last HgbA1c level categories | No Test | 4355.496 ± 20731.2 |
|  | <5.7 | 9993.659 ± 32218.04 |
|  | 5.7-6.4 | 14384.6 ± 38306.14 |
|  | 6.5-7.9 | 22676.88 ± 48046.23 |
|  | ≥8.0 | 34899.74 ± 59904.72 |

**Table 3:** Linear correlation (Spearman) between study variables and future medical cost.

| Variables | r | P |
| --- | --- | --- |
| Mean Age(years) | 0.291 | <0.001 |
| Male Gender | 0.005 | 0.672 |
| Mean number of Chronic Medications (N) | 0.3011 | 0.048 |
| Mean number of outpatient visits/Year (N) | 0. 259 | 0.036 |
| Mean number of Hospitalizations /Year (N) | 0.1614 | 0.041 |
| Mean number of Hospital Days /Year (N) | 0.1495 | 0.046 |
| Mean number of ED admissions/Year (N) | 0.1214 | 0.034 |
| Mean last HgbA1c level (%) | 0.1943 | 0.028 |
| Mean last Hemoglobin level (g/dL) | 0.01 | 0.321 |
| Mean last Creatinine level (mg/dL) | 0.102 | 0.058 |

In Table 3 we described correlation between the dependent variable and other variables selected into the regression model, in order to build the final score. Variables for which correlation was not statistically significant (gender, mean last creatinine, mean last hemoglobin level) or the association with future medical cost was obvious (e.g. age) were not included in the model. The number of hospital admissions per year and the number of hospitalization days per year showed co-linearity and contributed to cost prediction in a similar and significant manner. We decided to use only the *number* of hospital admissions per year because this indicator better reflects the dynamics of the patients and the intervention necessary for their care.

Blood glycosylated hemoglobin (HgA1c) level was found to be a strong indicator of future costs, even when its results were in the normal range (less than 5.5%).

After the first regression model (**Table 4**) identified the most significant variables, we incorporated dummy variables in our second regression model (**Table 5**), to achieve better prediction for a future medical cost model.

**Table 4:** Baseline model for future medical cost prediction.

| Variables | Coefficient | Standard Err. | 95% CI | P-value |
| --- | --- | --- | --- | --- |
| Mean number of ED admissions/Year (N) | 958.58 | 61.83 | 837.41-1079.76 | <0.001 |
| Mean number of outpatient visits/Year (N) | 211.69 | 4.39 | 203.08-220.30 | <0.001 |
| Mean number of Hospitalizations /Year (N) | 6,720.33 | 126.30 | 6472.79-6967.88 | <0.001 |
| Mean number of Chronic Medications (N) | 2,461.32 | 18.41 | 2425.23-2497.41 | <0.001 |
| Mean last HgbA1c level (%) | 496.86 | 14.94 | 467.58-526.13 | <0.001 |
| Intercept Cons. | 1,655.05 | 33.95 | 1588.50-1721.59 | <0.001 |

**Table 5:** Model for future medical cost prediction using dummy variables.

| Variables | Coefficient | Standard Err. | 95% CI | P-value |
| --- | --- | --- | --- | --- |
| Dummy Yes/No ED admissions/Year | 411.66 | 122.37 | 171.83-651.51 | <0.001 |
| Dummy- more than one ED admissions/Year | 878.37 | 182.64 | 520.39-1236.34 | <0.001 |
| Number of outpatient visits(0-30)/Year | 225.08 | 4.85 | 215.08-234.64 | <0.001 |
| Number of outpatient visits(30-50)/Year | 243.65 | 24.77 | 195.10-292.21 | <0.001 |
| Dummy- one Hospitalization /Year | 6,567.61 | 126.30 | 6235.89-6899.33 | <0.001 |
| Dummy- two Hospitalizations /Year | 13,913.11 | 439.67 | 13051-14774.85 | <0.001 |
| Dummy- three and more Hospitalizations/Year | 18,454.33 | 931.79 | 16628.03-20280.62 | <0.001 |
| Number of Chronic Medications | 2,361.32 | 19.41 | 2324.49-2399.41 | <0.001 |
| Dummy- HgbA1c No/Yes (less than 5.5%) | 323.86 | 114.69 | 99.07-548.66 | <0.001 |
| Dummy- HgbA1c Yes (5.5%-7.9%) | 2,542.93 | 119.66 | 2308.39-2777.46 | <0.001 |
| Dummy- HgbA1c Yes (8% and more) | 3,866.61 | 172.55 | 3528.41-4204.81 | <0.001 |
| Intercept Cons. | 1,688.04 | 34.88 | 1619.67-1756.41 | <0.001 |

In our dataset, 5,900 patients had uncontrolled diabetes mellitus (DM), according to their last measured HgA1c (>8%). We hypothesized and examined whether there was an independent and statistically significant synergistic relationship in the *combination* of uncontrolled DM with the other indicators using dummy variables. We found the association to be statistically significant with emergency department (ED) visits (1173 patients), hospitalizations (851 patients) and having *less* than 4 chronic medications (2164 patients), but not for outpatient visits (**Table 6**). To reflect this in our final score, a “penalty” was set for the relevant variables.

**Table 6:** Baseline predictive cost model using “penalty” dummy variables.

| Variables | Coefficient | Standard Err. | 95% CI | P-value |
| --- | --- | --- | --- | --- |
| Dummy- Yes/No ED admissions/Year | 417.66 | 122.34 | 177.83-657.07 | <0.001 |
| Dummy- more than one ED admissions/Year | 831.77 | 183.54 | 473.39-1196.34 | <0.001 |
| Dummy- ED admissions & Uncontrolled DM | 2,376.86 | 650.18 | 1102.53-3651.07 | <0.001 |
| Number of outpatient visits(0-30)/ Year | 226.34 | 4.88 | 216.88-236.64 | <0.001 |
| Number of outpatient visits(30-50)/ Year | 231.65 | 24.79 | 183.14-280.42 | <0.001 |
| Dummy- one Hospitalization /Year | 6,176.93 | 171.73 | 5840.89-6513.53 | <0.001 |
| Dummy- two Hospitalizations /Year | 13,349.84 | 441.67 | 12484-14284.85 | <0.001 |
| Dummy- three and more Hospitalizations/Year | 17,729.33 | 933.45 | 15618.03-19807.62 | <0.001 |
| Dummy- Hospitalizations & Uncontrolled DM | 10,224.45 | 789.37 | 8677.31-11771.6 | <0.001 |
| Number of Chronic Medications | 2,372.26 | 19.43 | 2334.49-2410.41 | <0.001 |
| Dummy- No Chronic Medications & Uncontrolled DM | 1,064.31 | 519.85 | 45.41-2083.21 | 0.041 |
| Dummy- HgbA1c No/Yes (less than 5.5%) | 433.18 | 115.16 | 207.07-658.66 | <0.001 |
| Dummy- HgbA1c Yes (5.5%-7.9%) | 2,234.33 | 125.66 | 1988.61-2479.86 | <0.001 |
| Dummy- HgbA1c Yes (8% and more) | 3,168.99 | 190.81 | 2794.95-3542.81 | <0.001 |
| Intercept Cons. | 1,693.31 | 34.88 | 1624.93-1761.69 | <0.001 |

The final predictive model was formulated using the following indicators:

- The mean number of chronic disease medications purchased per year. This was the most significant indicator. Due to a low number of patients (270), those receiving 16 medications or higher were grouped together.
- The mean number of outpatient visits per year. Up to 30 visits per year showed good correlation. 30-50 visits per year were still significant, but each additional visit had less influence on overall future costs. More than 50 visits per year (1300) were excluded from the final calculation.
- The mean number of ED admissions per year. The first visit was the most significant.
- The mean number of hospital admissions per year. Up to 3 visits a year showed very significant financial correlation. More than 4 visits failed to increase the risk and were therefore grouped with 3 visits per year.
- The last measured HgA1c level.
- The aforementioned “penalties” for uncontrolled DM concurrent with other variables.

## DISCUSSION

Leading global health organizations invest considerable resources in defining the requirements for pro-active intervention programs aimed at reducing healthcare expenditure by slowing or stopping the deterioration of patients at risk of becoming High Need-High Cost [22-23]. Such intervention plans require the timely identification of the appropriate target patient population.

Whereas health care systems currently tend to rely on the intuitive detection of “complex patients” by family physicians (qualitative detection) [18, 23] or the use of computerized detection based solely on the number of chronic diseases (quantitative detection) [24-25], these methods have proven ineffective, and allow many of those in need to go unnoticed while including patients that will stay stable for many years.

There is a noticeable increase in the use of predictive models, using linear regression techniques, to estimate the expected total health care costs for managed care organizations. Various data sources have been used for such predictive modeling, including medical and pharmaceutical insurance claims, health risk-assessment surveys, and laboratory data [26-28]. The computerized predictive models can be used for proactive identification of high-risk patients for the effective adaptation of disease management programs [28-31].

In this study, we developed a predictive model, which is based on numerical indicators with the most significant association with patient medical expenditure within 5 years, as an expression of health deterioration during this time period. We hypothesize that this model will enable us to detect a population at risk for deterioration, with a sufficient time period for effective pro-active intervention in primary care.

The next goal will be to develop a weighted numerical risk score, based on the relative influence of the variables, included in our predictive model.

A numerical point system awarded to each of these variables, according to its appropriate risk value, will be calculated into a final predictive score between 0-100. A higher score will indicate a higher risk for health deterioration within 5 years.

This calculated risk score will enable us to build a mechanism for the efficient identification of complex patients, and to put additional focus on their management in an attempt to prevent deterioration to HNHC, as well as to evaluate the effectiveness of specific interventions.

The indicators introduced into this calculated risk will guide healthcare providers to the needed areas of intervention. The display of indicators also promotes optimization of care management and continuity of care.

The introduction of this calculated risk score for deterioration is expected to focus the attention of primary care teams on the population that will benefit most from it. It is important to continually evaluate each intervention while receiving feedback from family physicians who are familiar with their “complex” patients, know patients’ ability to manage their illness and the factors which affect their disease control. Such feedback is very important because effective involvement of the primary care team is critical for the success of the intervention [18-19].

### Strengths and weaknesses

The main strengths of the study are its large size, and its real-world, population-based nature. An additional strength is the inclusion of a multitude of numerical variables that could be identified in most health care organizations.

However, there are some weaknesses, including its retrospective design. Furthermore, relevant clinical variables such as the specific diagnosis of a chronic illness (both somatic and psychiatric) and the severity of its presentation were not included in the model because this data was not accurately presented in the medical records.

In conclusion, we formulated a predictive model for precise and early identification of patients whose prognosis may be altered before they experience medical deterioration, by using numerical indicators existing in standard EMR data. Using this model may benefit the population that will potentially improve considerably following an intervention program.

The next step should be to integrate a calculated risk score for health deterioration into the LHS EMR and to prospectively evaluate its contribution towards reducing the utilization of health care resources and possible deterioration of patients to HNHC.

## Data Availability

All data produced in the present study are available upon reasonable request to the authors

## Acknowledgments

authors would like to express their deep and sincere gratitude to Mr. Haim Fernandes, Mr. Ilia Merhasin, and Mrs. Etty Cohen-Adar.

## Notes

### Competing Interest Statement

The authors have declared no competing interest.

### Funding Statement

This study did not receive any funding

### Author Declarations

The study protocol was approved by the Shamir Medical Center Review Board and the Research Committee of LHS ( ID number: 0235-18-ASAF, Date: 9.10.2018)

